# Adaptive mobility responses during Hurricanes Helene and Milton in 2024

**DOI:** 10.1101/2025.07.02.25330752

**Authors:** Qing Yao, Victoria D. Lynch, Molei Liu, Xiao Wu, Robbie M. Parks, Sen Pei

**Affiliations:** Department of Environmental Health Sciences, Mailman School of Public Health, Columbia University, New York, NY, USA; Department of Biostatistics, Mailman School of Public Health, Columbia University, New York, NY, USA; Department of Biostatistics, Peking University Health Science Center, Beijing, China; Beijing International Center for Mathematical Research, Peking University, Beijing, China

**Keywords:** human mobility, hurricane, climate extremes, adaptation, flood risk

## Abstract

Adaptation is crucial for minimizing the societal impacts of tropical cyclones amid climate change. Using 3.56 billion high-resolution foot-traffic records from mobile devices, we analyzed human mobility patterns during Hurricanes Helene and Milton, which struck the southeastern United States in 2024. We observed marked differences in adaptive mobility responses across geographic regions with varying levels of historical hurricane exposure. Milton primarily impacted coastal areas with frequent hurricane exposure and prompted sharp increases in out-region travel prior to landfall and sustained elevated mobility in the post-disaster period. In contrast, Helene affected mostly inland areas, where mobility changes were modest and largely within natural variation. Within Helene-affected regions, coastal counties showed stronger mobility responses than inland counties. Our findings underscore the need for tailoring disaster preparedness and response strategies to the specific characteristics of affected populations.

## 1. Introduction

Climate change has profoundly affected human societies, notably through the increased frequency and severity of natural disasters that endanger public health and disrupt economic activity. Understanding human mobility during extreme weather events is essential for effective disaster preparedness and climate adaptation. Hurricanes and other tropical cyclones are among the most harmful of these events with devastating effects on many health outcomes [1,2], but rigorously quantifying their impact on health is complicated by human mobility before, during, and after landfalls. Accurately measuring population movement is crucial for quantifying community adaptations and assessing the influence of tropical cyclones on health [3], yet this task remains challenging in post-disaster environments [4]. The effect of tropical cyclones on population movement can vary by sociodemographic, geographic, or infrastructure-related factors. Evacuation during tropical cyclones differs by race and income [5], with wealthy white residents more likely to evacuate ahead of hurricanes than low-income, Black, or Hispanic residents [6,7]. Transportation infrastructure, particularly related to car traffic, also shapes the decision and ability to evacuate [8,9]. Beyond structural factors, individual and social dynamics – such as experience, risk perception, and social norms – can influence adaptive behaviors related to tropical cyclones, though most research has focused on disaster preparedness rather than mobility [10,11].

The availability of high-resolution human mobility data derived from mobile phones presents an opportunity to study population movement during climate-related disasters with unprecedented detail. Analyses of these mobility data have provided valuable insights into evacuation behaviors [12], adaptive responses [13], recovery dynamics [13,14], prediction of mobility and trajectories [13,15], and mobility network changes [16] during hurricanes and other climate-related disasters [6,17–19]. While this growing body of work has advanced our understanding of hurricane-related mobility patterns, most studies focus on single events, typically coastal regions with high historical exposure. Research on tropical cyclones has not systematically compared adaptive mobility responses across populations with varying historical hurricane exposure. As climate change expands the geographical range of tropical cyclones into previously unaffected areas, this gap limits our understanding of the unique needs of different populations to tailor disaster preparedness and response strategies. In particular, the adaptive mobility of inland populations – who have lower hurricane exposure and risk perception than coastal populations – remains understudied [20].

In 2024, two major hurricanes, Helene and Milton, successively hit the southeastern United States (US). On September 26th, Helene (Category 4 at both its strongest and landfall) made landfall near Perry, Florida before moving inland where it led to historic rainfall and flooding in Georgia, South Carolina, North Carolina, Tennessee and Virginia, resulting in an estimated 219 deaths and losses of $78.7 billion [21]. On October 9th, Milton (Category 5 at its strongest and Category 3 at landfall) made landfall near Siesta Key, Florida, and passed through central Florida, resulting in an estimated 32 deaths and losses of $34.3 billion [21]. The devastating impacts of these two hurricanes provide an opportunity to examine population dynamics across coastal and inland areas, including low-exposure regions that are often underrepresented in hurricane mobility studies. Because they struck the southeastern United States within weeks of each other, our study design minimizes geographic and temporal confounders. This comparison highlights the contrasting responses of inland and coastal populations and offers insights to strengthen future preparedness in both affected inland and coastal areas [22].

## 2. Method

### 2.1 Mobility data

Here, we analyzed high-resolution foot-traffic data in the US to evaluate changes in mobility in the weeks before, during, and after hurricane landfalls. The foot-traffic data were provided by Advan [23], a company that collected human visitation data derived from mobile devices using all major U.S. mobile carriers (e.g., Verizon, T-Mobile, AT&T). The anonymous mobility dataset reports de-identified daily visits to points of interest (POIs, e.g., restaurants, grocery stores, etc.) for a random sample of cellphone users in the US and users’ home locations at the census block group level. According to Advan’s documentation, only device panels with explicit permission are used. Visits were identified when a device entered the defined polygons of POIs [24]. We used the normalized visit field, which represents raw visit counts scaled using the mobile device sampling rate for the state in which the POI is located. This scaling helps adjust for differences in device panel sizes across states and mitigates potential biases due to uneven geographic or carrier representation. During the week of September 9–15, 2024, for example, the dataset included approximately 835.0 million raw visits to 5.565 million POIs, corresponding to about 656.6 million aggregated “raw visitors,” which scaled to about 10.80 billion normalized visits nationally.

We aggregated the foot-traffic data from the census block groups and POI levels to the county level and created a daily county-level mobility matrix {*M*_*ij,t*_} across all 3,144 counties in the US from September 9th, 2024 to October 20th, 2024. Here *M*_*ij,t*_ denotes the daily number of visits from county to *j* on day *t*.

### 2.2 Within- and out-region mobility

We defined the affected regions as the counties whose geographic centers were within a specified distance, *d*, of the storm track, denoted by *R*_*d*_ [25] (see Supplementary Materials, Fig. S1). The distance to the storm track was measured as the shortest distance between a county’s centroid and the storm’s path. Counties outside the defined affected region are considered ‘outside’. ‘Within-region’ visits refer to visits where the origin ‘home’ and destination POIs are located inside the affected counties, capturing local activity within the impact area. ‘Out-region’ visits denote visits originating inside the affected counties but ending at POIs outside them, capturing travel that leaves or evacuates the impacted area. Both are adjusted for the sampling rates of mobile devices [24]. For each day *t*, the total number of within-region visits was calculated as the sum of all visits from each affected county to any counties within 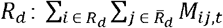. The total number of out-region visits on day *t* was calculated as the sum of visits from any affected counties in *R*_*d*_ to counties outside 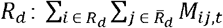 where 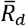 denotes the set of counties outside the hurricane-affected area *R*_*d*_. To define a list of destination counties that received the majority of visits from hurricane-affected areas, we computed the out-region visits to all other counties during the study period and selected the counties that cumulatively received over 95% of the total out-region visits. The travel distance of visitors from an affected county to a destination county *j* was measured as the geodesic distance between their centroids. The average travel distance of out-region visits was weighted by the number of visits between affected and destination counties.

### 2.3 Mobility changes compared to the baseline

To establish the counterfactual mobility patterns without the disruption of hurricanes, we chose the week of September 9th, 2024, two weeks prior to the landfall week of Helene, as the baseline period for both hurricanes (Fig. 1). For Milton, this earlier baseline was necessary because the weeks prior to its landfall (October 9th) overlapped with the impact period of Helene. Mobility during this period was already abnormal and may not represent the normal condition for Milton-affected population (Fig. 1). Using the week of September 9th ensures a more stable reference point for comparison. To account for the mobility variation within a week (e.g., weekday versus weekend), we matched the day of week when computing mobility changes compared to the baseline week. To assess potential seasonal effects, we repeated the analysis for the same calendar weeks in 2023, a year without hurricanes (Fig. S2). Results show smaller mobility fluctuations relative to baseline week of 2023 and different patterns as 2024, reinforcing that the major mobility shifts in 2024 were driven by two hurricanes. We further used the foot-traffic data in 2023 to quantify the intrinsic mobility variation without the impact of hurricanes. To estimate the distributions of mobility changes for within- and out-region visits due to natural fluctuations, we examined historical mobility data from September 11th, 2023 to November 5th, 2023, matching the study period with hurricanes in 2024. For each region, daily out-region mobility changes relative to the average mobility on each day of the week during this period were computed. We employed a Gaussian kernel density estimation to estimate the distributions of mobility changes, which informed the 95% CIs (confidence intervals) of natural mobility fluctuations. P-values of observed mobility changes were computed using the estimated distributions of mobility variation. The 95% CIs of the intrinsic variation of mobility to each destination county was computed in the same way using the number of visits from hurricane-affected areas. Mobility changes in 2024 outside the 95% CIs of these distributions were deemed significantly different from the baseline.

**Fig. 1.**
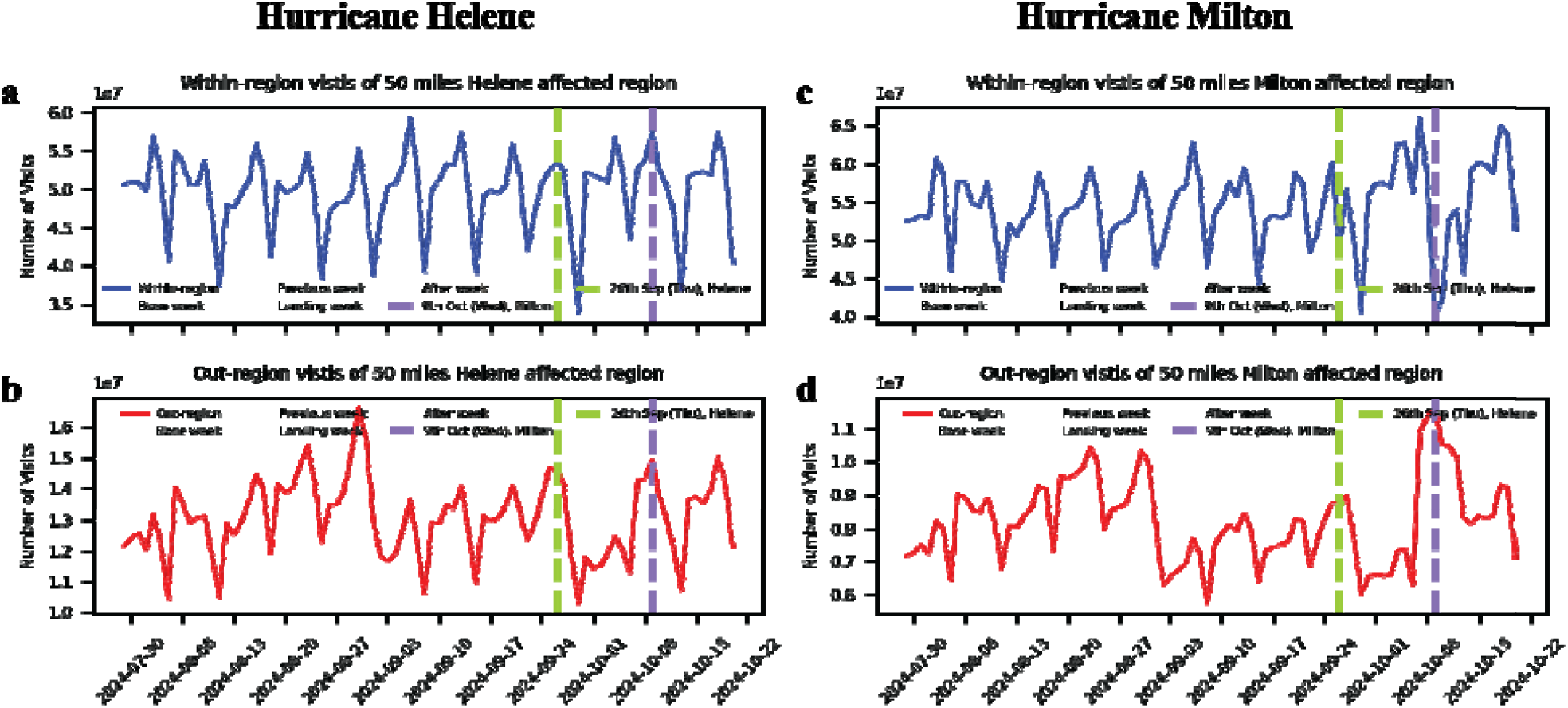
Time series of POI visits for regions affected by hurricanes Milton and Helene. The affected regions were defined as the counties whose geographical centers were within 50 miles of the storm track. **a**. Daily number of visits to POIs within the affected region for Helene. The four shaded areas show the baseline week (grey), the week before hurricane landfall (orange), the landfall week (teal), and the week after the landfall (light blue), respectively (from left to right). The green vertical dash line shows the landfall date of Helene. The purple vertical dash line shows the landfall date of Milton. **b**. Daily number of visits to POIs outside the affected region for Helene. **c**. Daily number of visits to POIs within the affected region for Milton. **d**. Daily number of visits to POIs outside the affected region for Milton.

## 3. Results

### 3.1 Overall mobility before, during, and after hurricane landfalls

We identified 3.56 billion POI visits from sampled mobile phone users within 50 miles (∼80.5 km) of the storm tracks during the study period (September 9th, 2024 to October 20th, 2024), representing the mobility of 24.4 million residents in affected regions for both hurricanes. For Hurricane Helene, residents from 271 affected counties (mostly inland) visited 230 out-region counties, while for Hurricane Milton, residents from 21 affected counties visited 310 out-region counties. Additional analyses for other distances (10 to 100 miles) are available in the Supplementary Materials (Tab. S1, Fig. S3). Without the impact of hurricanes, both within-region and out-region visits—across different distance cutoffs—exhibited consistent weekly patterns, with lower activity on weekends and higher activity on weekdays (Fig 1, Fig. S3). However, these patterns were disrupted prior to the landing of hurricanes.

### 3.2 Comparison of adaptive mobility responses during Helene and Milton

For Hurricane Helene, out-region visits increased modestly by 5.1% during the three days before landfall despite emergency declarations and evacuation orders [26]. This out-region visit change was within the 95% CI of natural mobility variation [-7.7%, 8.0%] (Fig. S4). During the same period, within-region visits declined by 2.8%, falling inside the 95% CI of natural variation [-8.0%, 8.6%] (Fig. S4). In the four days following landfall, out-region visits remained near baseline levels with a slight 0.7% decrease (p>0.05, 95% CI [-7.7%, 8.0%]), while within-region visits dropped by 8.4% (p<0.05, 95% CI [-8.0%, 8.6%]). One week after landfall, out-region travel declined to 8.3% below baseline levels (p<0.05, 95% CI [-7.7%, 8.0%]) (Fig. 2a and 2b). Overall, mobility changes during the landfall week were mostly within or close to the range of natural mobility variation. Out-region travel during the post-landfall week fell significantly below the baseline.

**Fig. 2.**
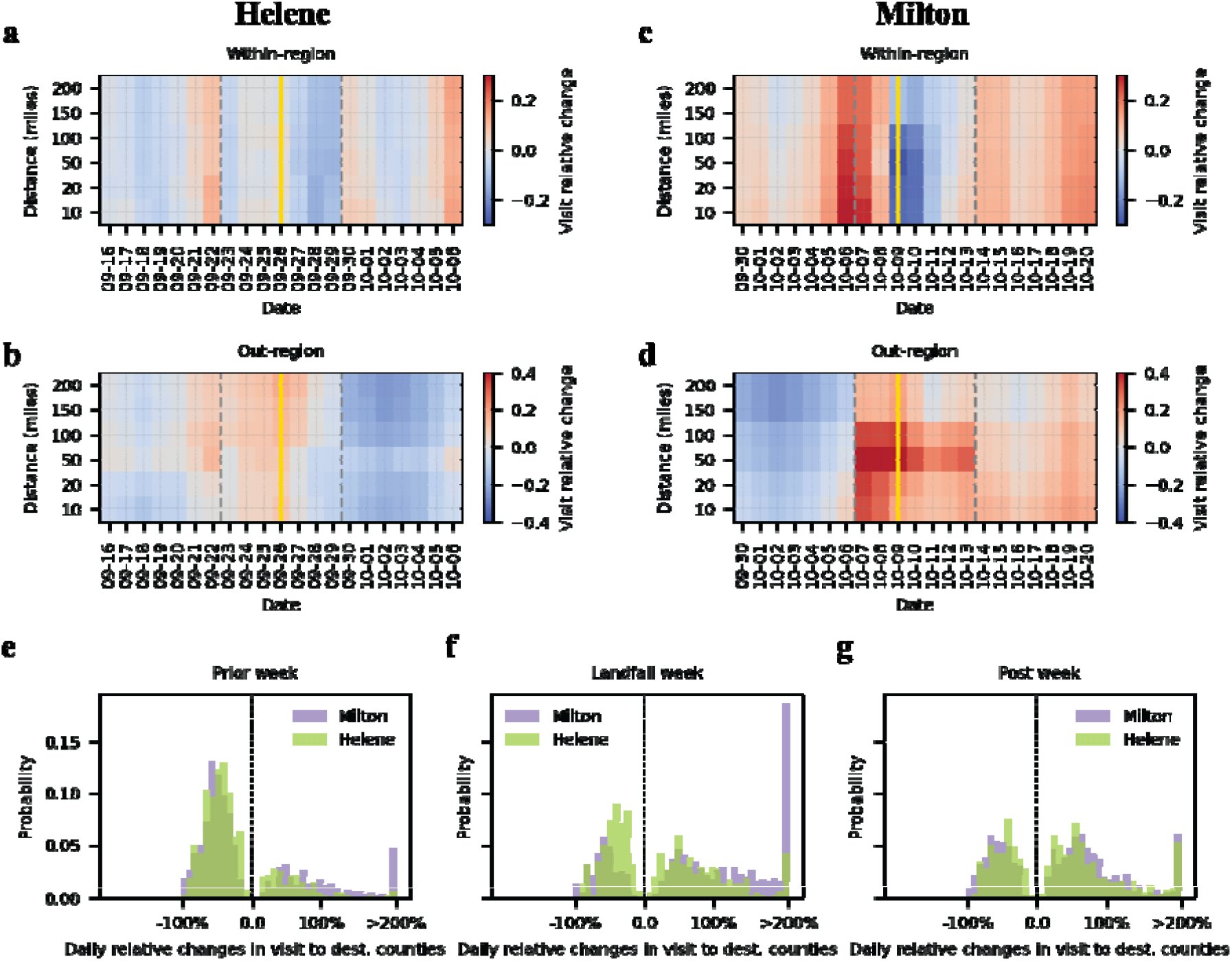
Temporal dynamics of relative mobility changes for hurricanes Helene and Milton. **a – d**. Each cell in the heatmaps represents the relative increase (red) or decrease (blue) in visit numbers compared to the baseline week (September 9^th^ – 15^th^, 2024). The yellow vertical lines indicate the landing dates and dashed grey lines show the landfall weeks. Each row represents the affected region defined using different distances to the storm track. **e**. For each destination county, we computed the daily relative changes in visit during the week prior to the landfall and selected the changes that were significantly different from the baseline (p<0.05). Distributions of these selected daily relative changes outside 95% CIs of natural variation are shown for Helene (green) and Milton (purple). **f – g**. Distributions of daily relative changes in single destination counties that were outside the 95% CI of natural variation are shown for the landfall week and the post-landfall week.

In contrast, Hurricane Milton led to a significant rise in both out-region (29.2%, p<0.05, 95% CI [-9.7%, 10.2%]) and within-region (18.7%, p<0.05, 95% CI [-10.1%, 10.3%]) visits beginning three days prior to the landfall, coinciding with the state of emergency declaration and one day before evacuation orders were issued [27]. On the landfall day, within-region visits dropped sharply by 29.6% (p<0.05, 95% CI [-10.1%, 10.3%]), while out-region visits surged 43.3% above baseline (p<0.05, 95% CI [-9.7%, 10.2%]), indicating large-scale migration (Fig. 2c and 2d). In the weekends of post-landfall week, within-region mobility rebounded, surpassing baseline levels (15.5%, p<0.05, 95% CI [-10.1%, 10.3%]), while out-region mobility remained higher than baseline (13.5%, p<0.05, 95% CI [-9.7%, 10.2%]). These shifts in mobility substantially exceeded the range of natural variability observed during the same period in the previous year (Fig. S4). This pattern was consistent across affected regions defined by different distances from the storm track (Fig. S3). To assess the robustness of our findings to different population sizes—we recalculated within- and out-region mobility changes through per-capita visit counts (visits divided by county population) and then aggregated the county-level relative changes to the affected region using the median, which reduces the influence of unusually large per-capita changes in a small number of counties (Fig. S5).

### 3.3 Mobility changes in single destination counties

We analyzed the relative percentage changes in out-region visits to each destination county. Due to the natural fluctuations of human mobility, visits to destination counties changed over time. To select mobility changes beyond natural fluctuations, we computed the 95% CI of historical mobility variation for each destination county and filtered the days with visits from the affected areas that were significantly different from the baseline (p<0.05). We compared the distributions of signficant mobility changes in the selected “destination county-days” for Helene and Milton in three weeks – the week prior to landfall, landfall week, and post-landfall week (Fig. 2e-g).

In the week before landfall, destination counties of the Milton-affected areas that experienced increased visits had on average a higher relative increase than those of the Helene-affected areas (Fig. 2e). Visits to many destination counties of the Milton-affected areas increased over 100%. In contrast, most increase of visits to the destination counties of the Helene-affected areas were within 100%. During the landfall week, out-region visit patterns showed greater variability in Milton-affected areas (Fig. 2f). Notably, 203 “destination county-days” had an increase of daily visits over 200% for Milton, while only 24 “destination county-days” experienced over 200% increase for Helene. In the post-landfall week, the distributions of significant mobility changes (both increase and decrease) in single destination counties became similar for Helene and Milton (Fig. 2g). This pattern remained similar for affected areas defined using different distances (Fig. S6).

We further examined the geographical patterns of out-region visits during the landfall week. Destination counties where daily mobility were significantly different from the baseline in at least one day are visualized (Fig. 3a and 3b). The cumulative changes of visits to these counties during the landfall week are presented. Residents from both hurricane-affected areas migrated to many non-neighboring counties across the US, including long-distance migrations beyond the immediate neighboring counties. During both landfalls, there was a considerable increase from affected counties in visits to densely populated areas in Florida and Southern California. The geographical relative out-region changes (Fig. S7) reveal contrasting response patterns between Helene and Milton. Milton triggered strong relative mobility increases both around affected areas and in several distant metropolitan regions, broadly consistent with the raw visit patterns. In contrast, Helene’s effects were more muted and concentrated outside its immediate neighboring counties. Those national view illustrate that large hurricanes may propagate disruptions across the wider mobility network, extending their influence beyond the directly affected counties. Focusing on the destination counties with increased visits, Milton led to a considerably higher volume of out-region visits, despite having 4.6 million fewer residents, and longer travel distances (Fig. 3c). The weighted average travel distance for visits to destination counties with increased mobility was 956.2 km for Milton compared to 418.9 km for Helene during the landfall week. For destination counties with decreased visits, Helene-affected regions experienced a more pronounced decline within 1,000 km, whereas Milton-affected regions showed declines at greater distances, primarily between 1,000 and 2,000 km (Fig. 2d). These findings were robust to the definition of affected areas using different distances (Figs. S8-S9).

**Fig. 3.**
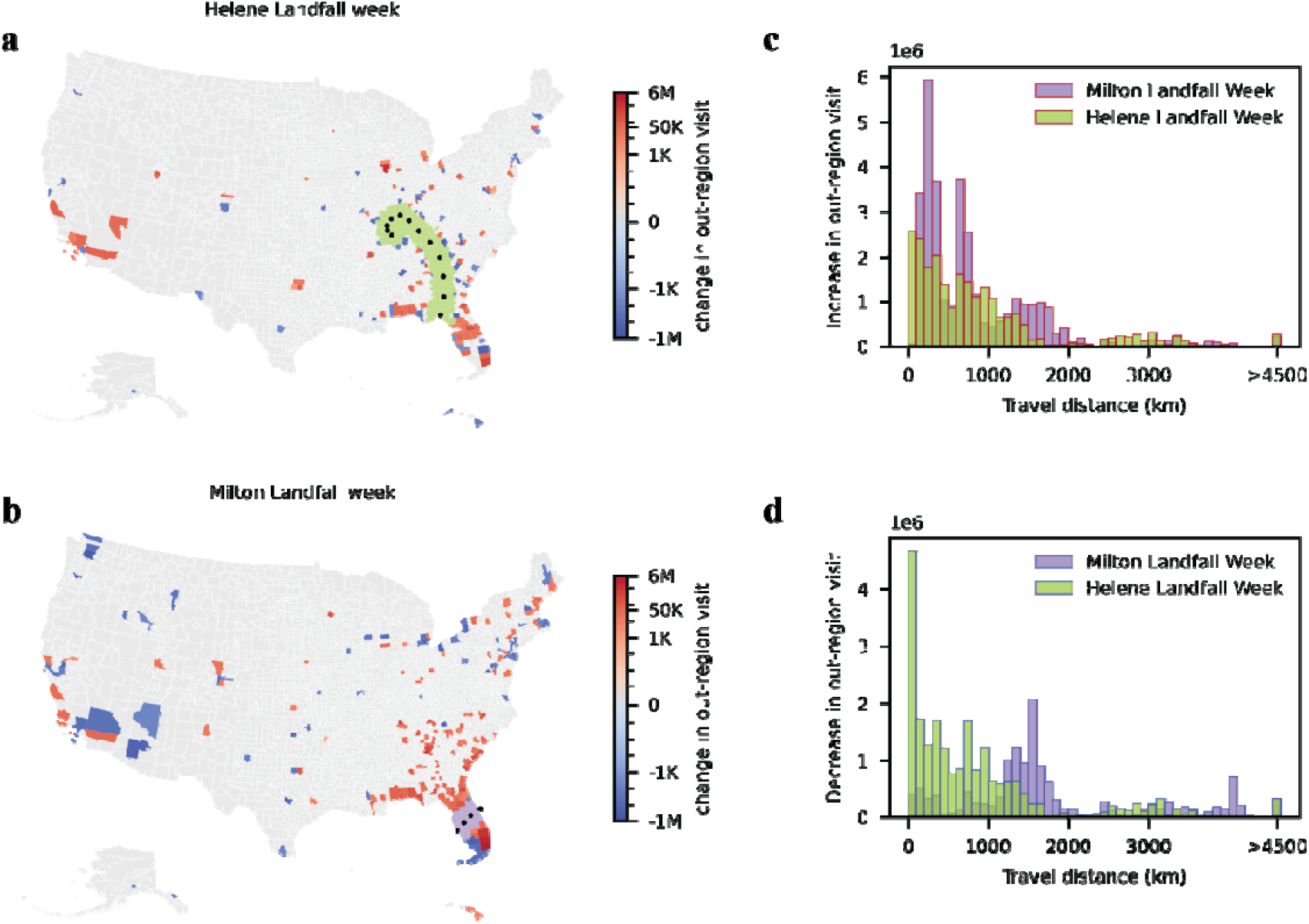
Population migration during the landfall weeks of hurricane Helene and Milton. **a – b**. Maps depict absolute changes in out-region visits to destination counties where the mobility was significantly different from the baseline in at least one day (p<0.05). Color shows the cumulative change in out-region visits during the landfall week. Red indicates increased visits while blue represents decreased visits from the affected regions. Black dots mark the storm tracks. Purple areas are the affected counties **c**. Distributions of travel distance for increased out-region visits to destination counties. Each bar in the histogram shows the number of increased visits within a certain range of travel distance. For instance, the highest bar represents that there were approximately 6 million more visits to out-region counties located 200 – 300 km from the Milton-affected regions compared to the baseline mobility. **d**. Distributions of travel distance for decreased out-region visits to destination counties. The highest bar represents that there were about 5 million fewer visits to out-region counties located 0 – 100 km from the Helene-affected regions.

### 3.4 Mobility changes in coastal and inland populations for Helene-affected counties

The Helene-affected region included both coastal and inland counties. To examine the adaptive mobility changes during one single hurricane, we analyzed mobility changes separately for coastal (within 50 miles of coastline) and inland populations (Fig. 4a). Using mobility data from the same period in 2023, we estimated the distributions of natural mobility variation for coastal and inland counties (Fig. 4b). We observed that visits from coastal regions exhibited a sharper surge in out-region mobility in the days preceding landfall, reaching up to a 20% increase (p<0.05), whereas inland areas showed only modest changes within the 95% CIs of natural fluctuations (Fig. 4c). Moreover, the weighted average travel distance associated with significantly increased mobility was substantially higher in coastal counties, suggesting longer evacuation or relocation trips (Fig. 4d).

**Fig. 4.**
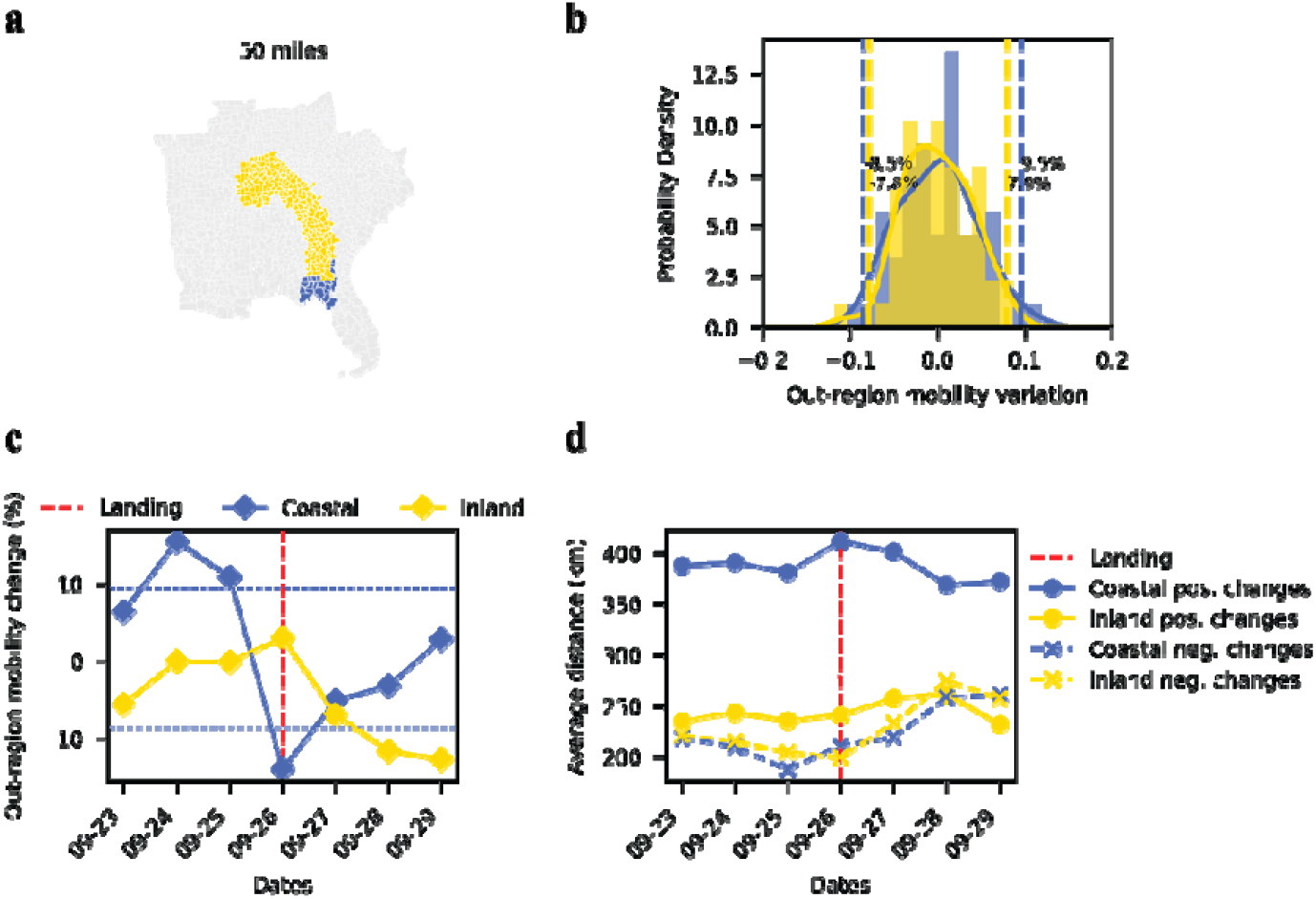
Comparison of mobility patterns between coastal and inland counties for the Helene-affected region. **a**. The coastal (yellow) and inland (blue) counties in the Helene-affected areas. **b**. Natural out-region mobility variations during the same period in the previous year. The 2.5th and 97.5th percentile thresholds are indicated by the vertical dashed lines. **c**. Time series of out-region mobility changes (%), with squares indicating changes in coastal (blue) and inland (yellow) counties. The horizontal dash lines show the 95% CIs of natural mobility variation in coastal and inland areas. **d**. Time series of weighted average travel distance (km) for positive (solid line with circles) and negative (dashed line with crosses) out-region visit changes.

Furthermore, we examined Federal Emergency Management Agency (FEMA) Major Disaster Declaration counties during Hurricane Helene [28]. Coastal counties with FEMA declarations and within the 50 mile storm track exhibited notably significantly higher out-region mobility changes compared with the natural variation in 2023, while inland areas showed more limited movement (Fig. S10).

## 4. Discussion

The contrast in mobility responses between the two hurricanes highlights the complexity of evacuation and adaptation behaviors. In the Milton-affected coastal region, residents exhibited prompt adaptive mobility, echoing previous research on hurricanes using mobility data, which emphasizes fast evacuation behaviours and recovery, even in unusual areas such as New York City [12–14], whereas those in the primarily inland Helene-affected areas showed more constrained mobility changes. These behavioral differences suggest that regions with greater experience in tropical cyclone-related evacuations may have a higher adaptive capacity.

Differential mobility responses may influence the causes of hurricane-related mortality. A post-disaster analysis revealed that most fatalities of Hurricane Helene occurred in counties with low historical hurricane risk, including many counties where we found limited adaptive mobility. In these inland areas, most deaths were due to river flooding, infrastructure damage, and indirect causes such as power outages and medical emergencies [29], in contrast to coastal communities where storm surges were the primary driver of fatality. State-level mortality data further underscore this inland–coastal distinction: freshwater flooding accounted for the majority of deaths in North Carolina and Tennessee, wind was predominant in Georgia, South Carolina, Virginia and Indiana, and storm surge and tornado-related fatalities were notable in Florida [30,31] (Tab. S2). The analysis illustrates the distinct threat posed by inland exposure, particularly in low hurricane risk areas.

Nevertheless, these differential responses and mortality risk cannot be attributed to a single factor. Instead, they are likely shaped by multiple interacting elements, including prior exposure to hurricane and flooding, differences in risk perceptions [20], socioeconomic status (SES) such as income and education [6,7], population density and social connectivity [32], disaster-resilient transportation infrastructures [33], and household preparedness levels [34]. In particular, higher SES and dense population in coastal areas, such as those affected by Milton, may provide greater access to transportation, information, and resources through concentrated infrastructure and tightly knit social networks, enabling faster evacuation and stronger pre-landfall mobility responses. In contrast, the more dispersed and lower-SES inland areas impacted by Helene likely lacked these supportive dynamics, potentially limiting their mobility responses. While storm intensity may also play a role, the greater pre-landfall movement during Milton could reflect heightened awareness and preparedness for a stronger storm or higher risk perception in areas with more frequent hurricane experience. Further investigation is needed to disentangle the relative contributions of these factors across geographic regions and to assess how different preparedness measures influence mobility patterns.

Our findings extend previous work on hurricane-related mobility by moving beyond single-event case studies and systematically comparing population responses across regions with contrasting historical hurricane exposure. The results revealed potential mobility constraints in inland areas compared to the coastal regions. This highlights the need for developing region-specific disaster response strategies that account for the complex interplay of these factors. As climate-related disasters become more frequent and risk perceptions shift in previously unaffected areas, proactive planning, targeted interventions, and concentrated resources toward the most relevant hazards in different region will be essential for enhancing resilience across diverse communities [35,36]. These heterogeneous responses also have implications for trajectory prediction models of evacuation, which often assume uniform adaptive behaviors. Incorporating differences in populations may improve forecasts of evacuation flows and inform preparedness planning.

There are important limitations inherent to using mobile device data. First, geographic variability in data density and restrictions on certain device panels may lead to uneven representation across regions. Additionally, signal disruptions during landfall and the exclusion of individuals without mobile access may introduce bias. However, focusing on relative changes in mobility helps mitigate the impact of imperfect sampling data on our results. Second, mobility patterns during the baseline week may not precisely represent counterfactual mobility during the study period in the absence of hurricanes. However, given the relative stability of weekly mobility patterns (Fig. S3), and the alternative baseline week analysis (August 5th – 11th, 2024), which produced consistent patterns with Fig. 2a-d (Fig. S11), the choice of baseline week is unlikely to substantially affect the qualitative findings. Lastly, the mobility of residents in the Milton-affected region may be influenced by the reporting of the devastating damages caused by Helene. However, our comparative analysis on the mobility of coastal and inland counties within the Helene-affected region yielded similar findings. Despite these limitations, this study provides a foundation for future research on population-specific mobility and behavioral responses to disasters and may inform studies on the social impacts of climate change.

## Data availability statement

The data that support the research are available in GitHub repository (https://github.com/Qing1011/hurricane_mobility).

## Acknowledgements

This study was supported by National Science Foundation (NSF) grant DMS-2229605 (Q.Y. and S.P.), National Institute of Environmental Health Sciences (NIEHS) grant T32 ES007322 (V.D.L.), NIEHS grant R00 ES033742 (V.D.L. and R.M.P.), NIEHS grant K01ES036202 (X.W.), NIEHS grant P30 ES009089 (V.D.L., R.M.P., and X.W.), and National Institute of Aging (NIA) grant P20 AG093975 (R.M.P. and X.W.).

