## Supplementary Materials for "Adaptive mobility responses during Hurricanes Helene and Milton in 2024"

**A. Storm tracks and affected regions**

We defined the affected regions as those counties within a given distance from the storm track (Fig. S1). The distance was measured between the storm track and the county’s geographical center. The distances are calculated on the projected coordinate reference system (EPSG:5070, the USA Contiguous Albers Equal Area CRS).

**B.** **Baseline week**

Figure S2 presents time series data spanning from July 29 to October 20, 2024, measured at three distances from the storm tracks (10 miles, 50 miles, and 100 miles). In the period without hurricanes, the data reveal consistent weekly patterns, which justified our choice to focus on comparisons spanning Monday to Sunday. Given the closeness in timing of the two hurricanes, we chose to use the same baseline week for both events to ensure consistency and comparability in our analysis.

**C** **Sensitivity analysis**

In the primary analysis the affected region was defined as counties within 50 miles of the storm track. Here, we performed the analysis where affected regions were defined at distances of 10 miles and 100 miles as a sensitivity analysis (Tab. S1). Closer regions (10 miles) experienced the most direct and intense storm impact. Intermediate regions (50 miles, the main analysis distance) exhibited a mix of responses, capturing both severe storm impact and voluntary mobility adjustments. And farther regions (100 miles) tend to experience more indirect effects, such as infrastructural disruptions, or secondary evacuations.

There are more destination counties for Helene at 10-mile distance regions, possibly due to the movement preferences and many small-area counties along the storm tracks. However, the contrasting pattern of mobility responses to the two hurricanes remained consistent in the following analysis.

**C.1 Relative mobility changes**

Closer regions (10 miles) exhibited the most substantial mobility shifts, with a broader distribution, while farther regions (100 miles) showed less pronounced variations (Fig. S3). The main analysis (50 miles) captured similar patterns, with Hurricane Milton associated with more extensive evacuation compared to the more constrained mobility observed in Hurricane Helene-affected regions, maintaining consistency across different distance thresholds.

**C.2 Heatmaps of changes in visits to destination counties compared to the baseline week**

Consistent with the main analysis, Hurricane Helene (Fig. S4 panel a) showed moderate out-region visit increases near its storm track and distant counties, while Hurricane Milton (Fig. S4 panel b) exhibited stronger increases across Florida and nearby metropolitan areas, with declines elsewhere. These mobility patterns remained consistent across distance thresholds.

**C.3 Histograms of visit changes across various travel distances**

The travel distance between affected county and destination county was measured as the geodesic distance between their centroids, using the WGS 84 geographic coordinate system (EPSG:4326).

Similar to the main analysis, Hurricane Helene showed more localized changes, with both increases and decreases concentrated within shorter distances. In contrast, Hurricane Milton exhibited a higher proportion of visit increases at shorter distances and more pronounced visit decreases at longer distances. Despite variations in magnitude, these travel patterns remained consistent across distance thresholds (Fig. S5).

**C.4 Mobility Variation within FEMA-Declared Disaster Counties**

We analyzed counties designated as FEMA Major Disaster Areas during Hurricane Helene. These counties were categorized into three groups: (i) coastal counties located within both 30- or 50-mile proximity to the coastline and within 50 miles of the storm track (blue); (ii) coastal counties outside the 50-mile storm track cutoff (olive); and (iii) all remaining inland counties (yellow) (Fig. S7a). Panels b and c show that coastal counties with FEMA declarations exhibited the most pronounced increases in out-region mobility prior to landfall, while inland counties without FEMA declarations displayed only minimal changes.

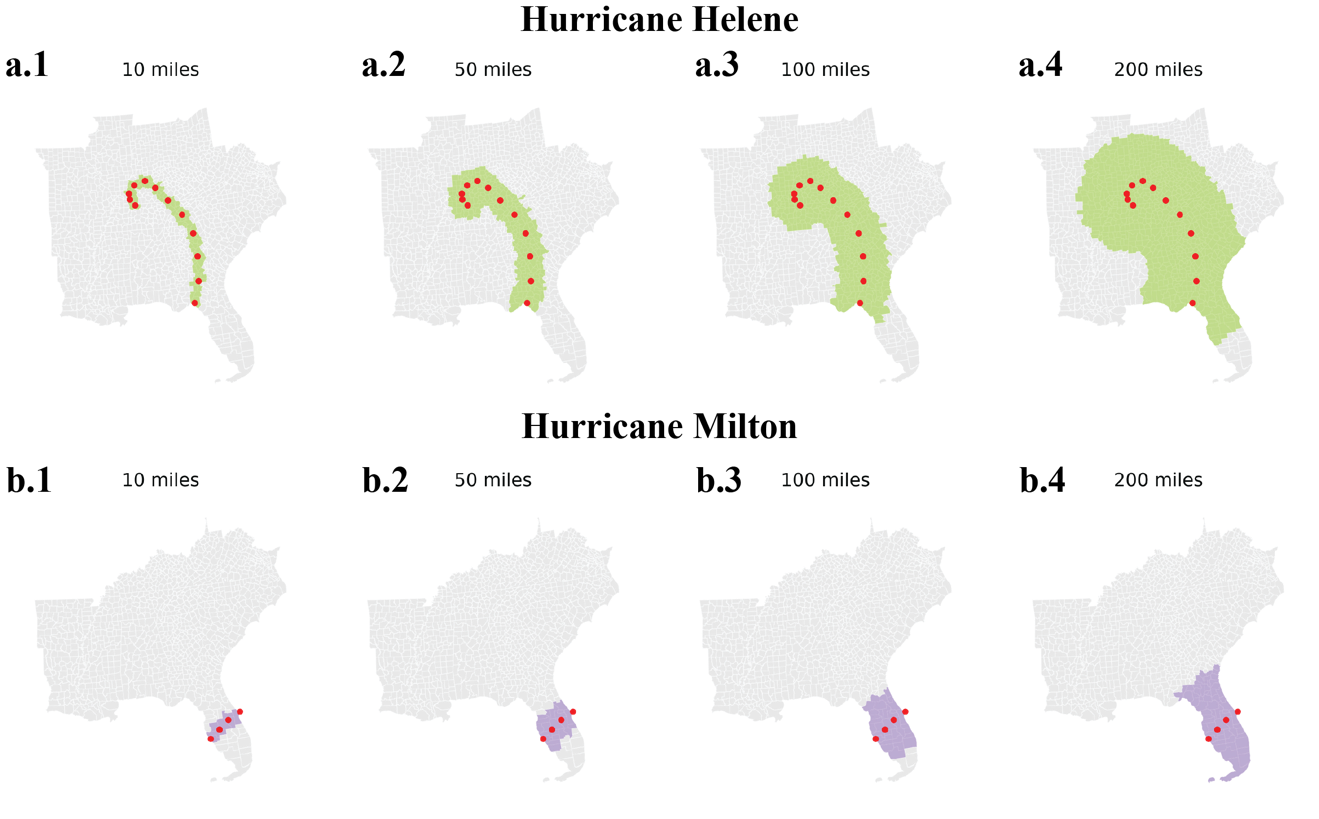

**Fig.S1 Storm tracks and the affected regions at different distances.** Sub-panels **a** and **b** illustrate the affected regions for Milton and Helene, respectively.

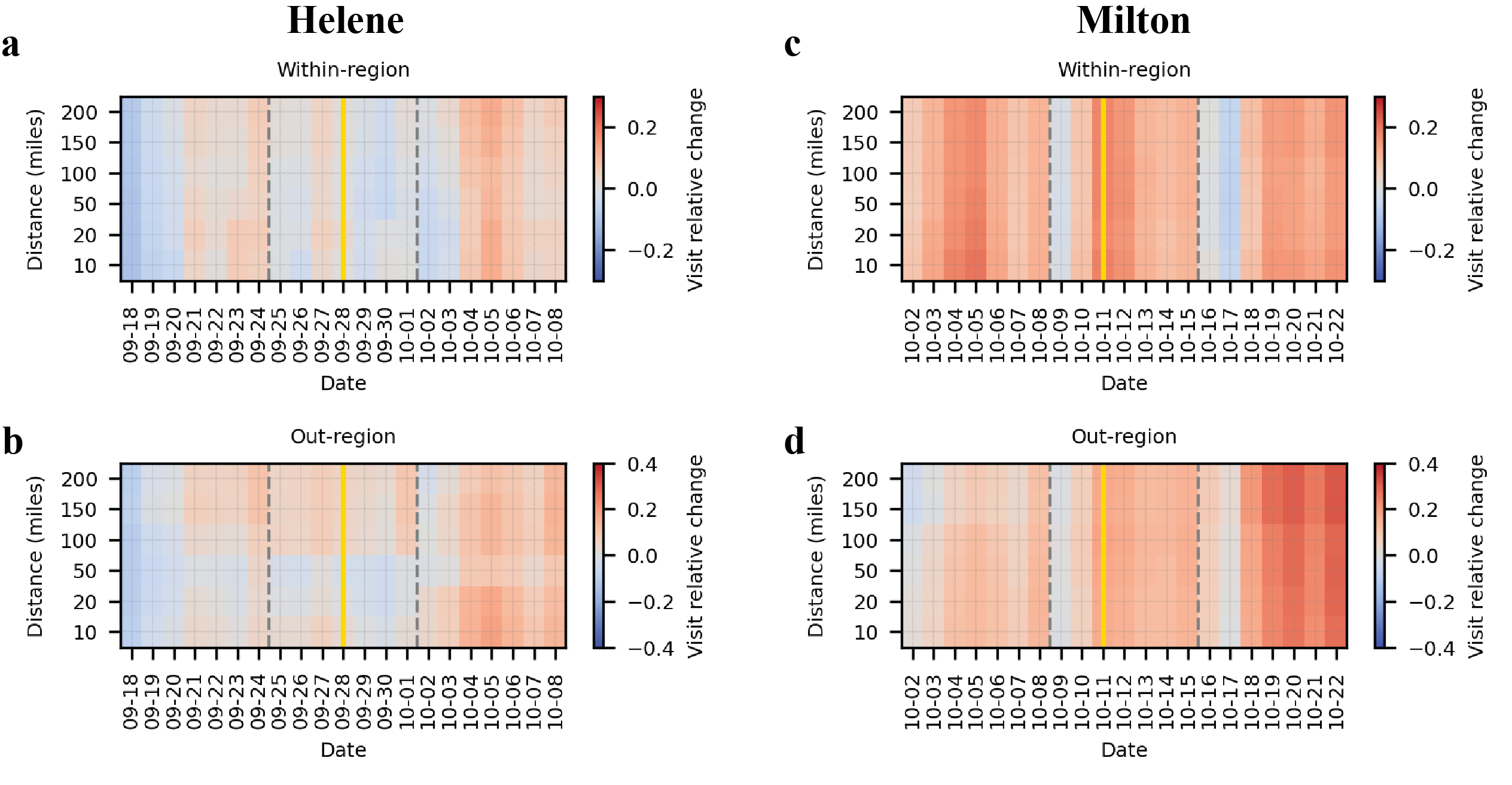

Fig. S2. Relative changes in within-region (a, c) and out-region (b, d) mobility during the same calendar weeks in 2023 (baseline week: September 11–17, 2023) corresponding to the periods of Hurricanes Helene (left panels) and Milton (right panels) in 2024. The yellow vertical lines mark the dates corresponding to the 2024 landfall days, and the dashed grey lines indicate the landfall weeks. Each row represents the affected region defined by different distances from the storm track. For the Helene-affected region, both within- and out-region mobility in 2023 show only minor fluctuations compared to baseline, with no major surges. For the Milton-affected region, most days exhibit moderate mobility increases. These patterns contrast with the pronounced mobility responses observed during the hurricanes in 2024.

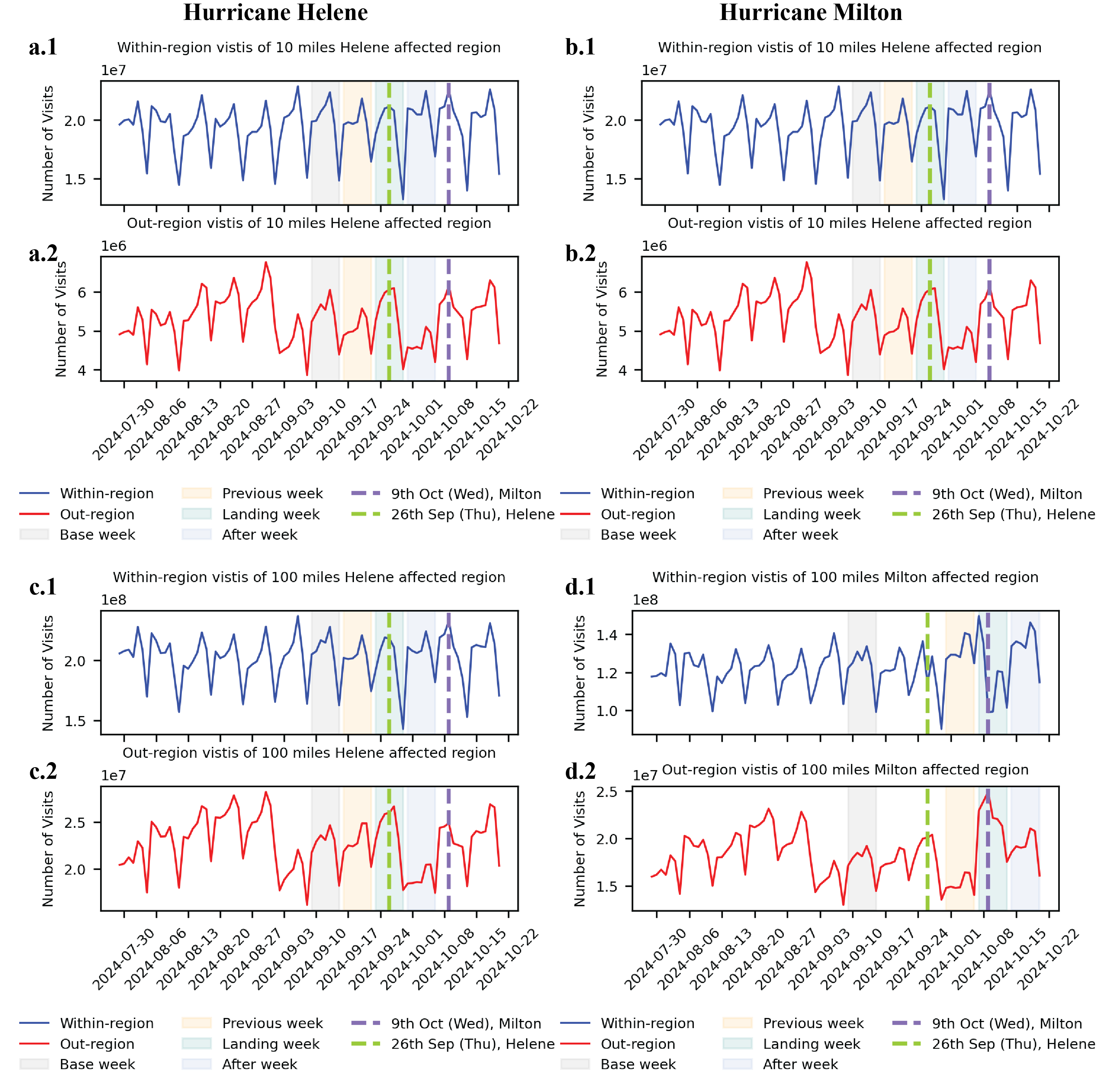

**Fig.S3 The time series of POI visits for hurricanes Helene and Milton.** Left column shows the results for Hurricane Helene and right columns displays the results for Hurricane Milton. All sub-panels **1** plot the within-region visits while all sub-panels **2** plot the out-region visits. The baseline week (grey), prior week (orange), landfall week (teal) and post week (blue) were shaded for the two hurricanes respectively. The dashed lines indicated the landing dates of the two hurricanes. Subpanels **a** and **b** show results for the 10-mile affected regions, while subpanels **c** and **d** correspond to the 100-mile affected regions.

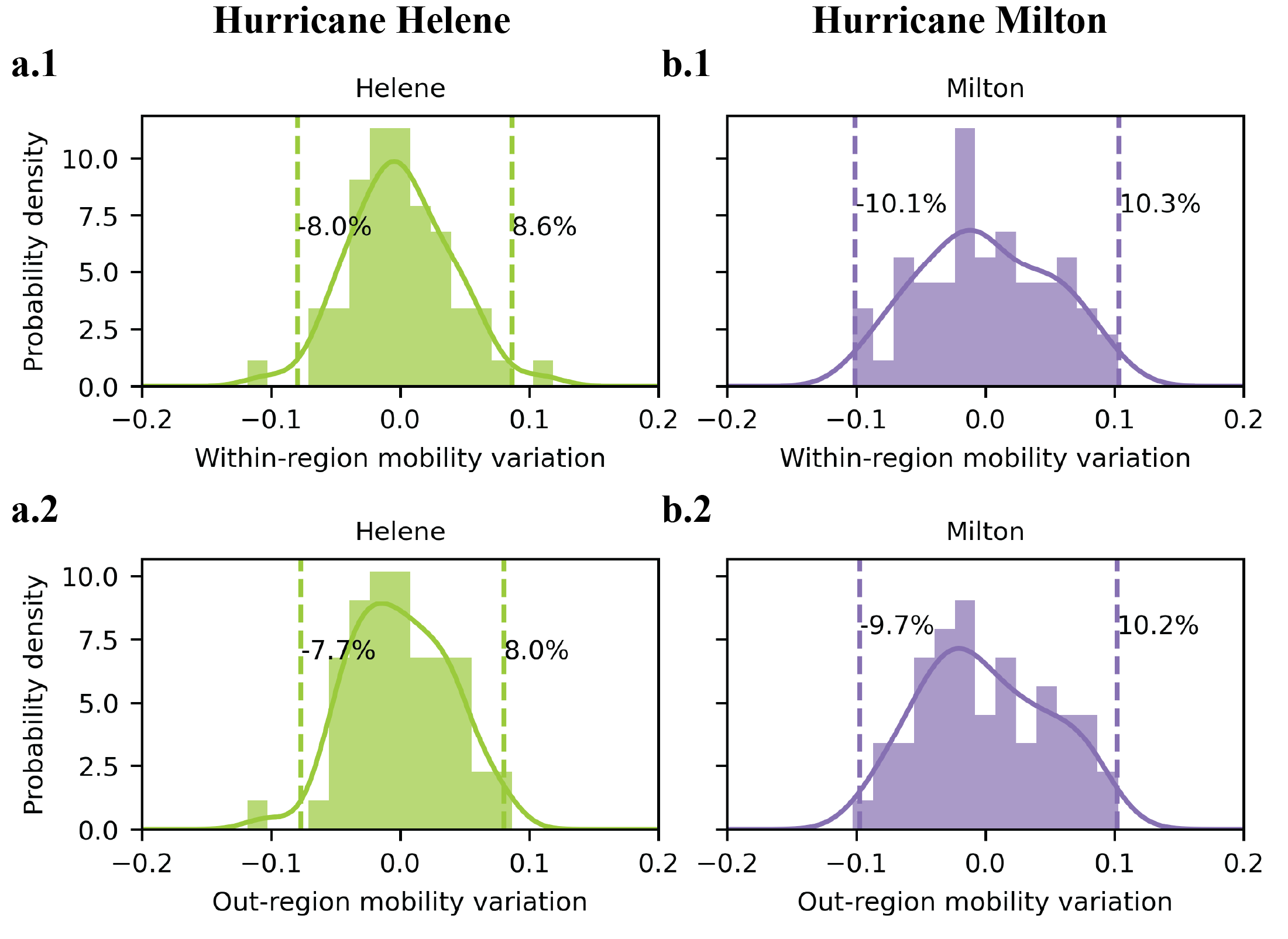

**Fig.S4 Natural mobility variations in the absence of hurricane events.** Panels **a** and **b** show the distribution of weekly mobility fluctuations within a 50-mile distance for regions affected by Hurricanes Helene and Milton, respectively. Panels **1** depict within-region mobility variation, while Panels **2** show out-region mobility variation. Dashed vertical lines indicate the 2.5th and 97.5th percentile thresholds, representing the range of natural variability.

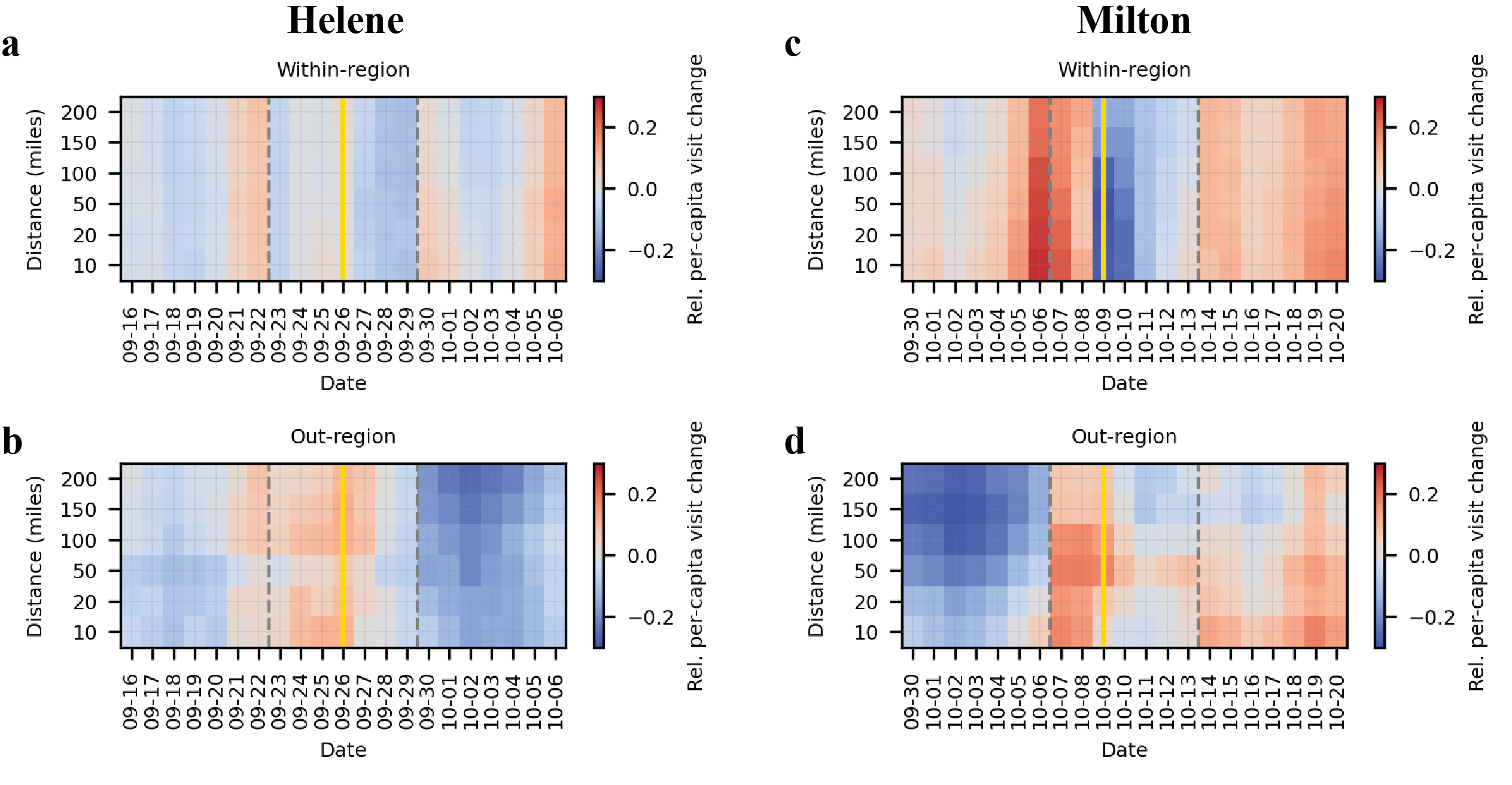

Fig. S5. Relative per-capita visit change by distance to storm track for within-region (top panels) and out-region (bottom panels) mobility during Hurricanes Helene (left) and Milton (right). Per-capita visit counts were computed using county population, and relative changes were calculated with respect to the baseline week and aggregated to affected regions using median. Patterns are consistent with the main results (Fig. 2), indicating that population size differences do not alter the qualitative findings.

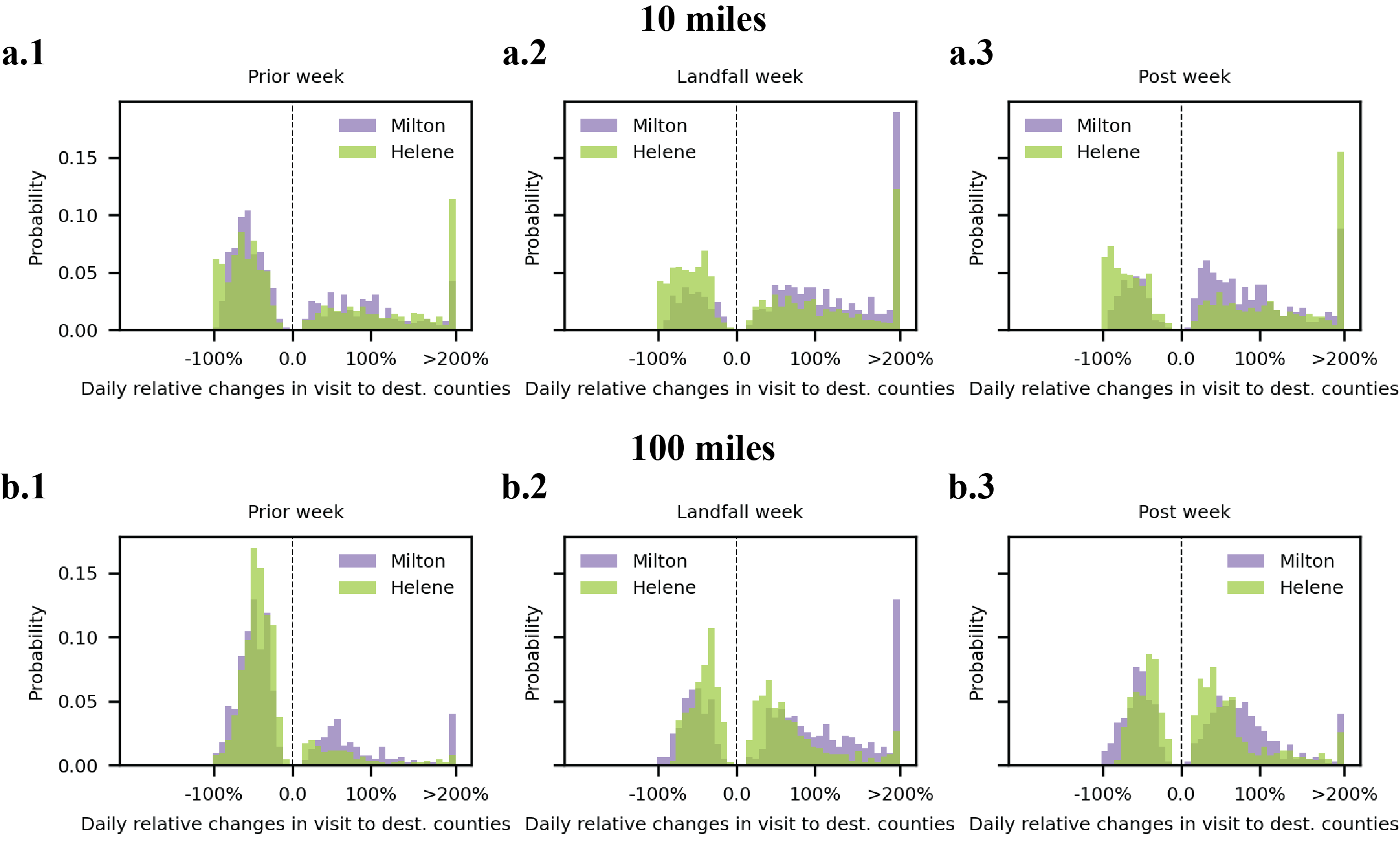

**Fig.S6 Daily relative mobility changes.** Sub**-**panels **a** and **b** plot the normalized distribution for the daily relative changes in visit to destination counties which exhibited significantly increase or decrease for hurricane-affected regions defined using the distance thresholds of 10 miles (**a**) and 100 miles (**b**).

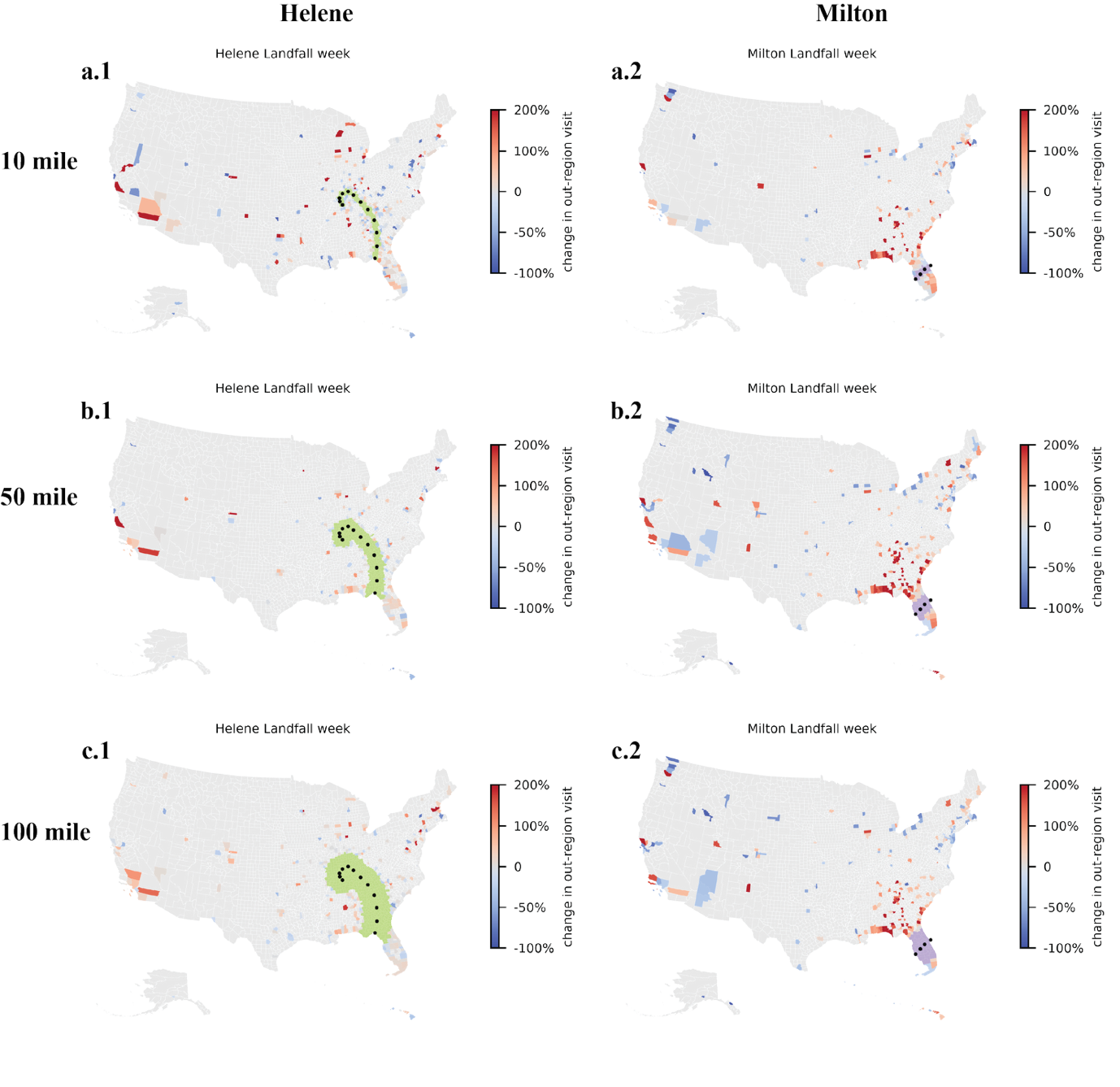

Fig. S7. Spatial distribution of relative changes in out-region visits during baseline and landfall weeks for Hurricanes Helene (left panels) and Milton (right panels). Panels a, b, and c correspond to affected regions defined by 10, 50, and 100 miles from the storm track, respectively.

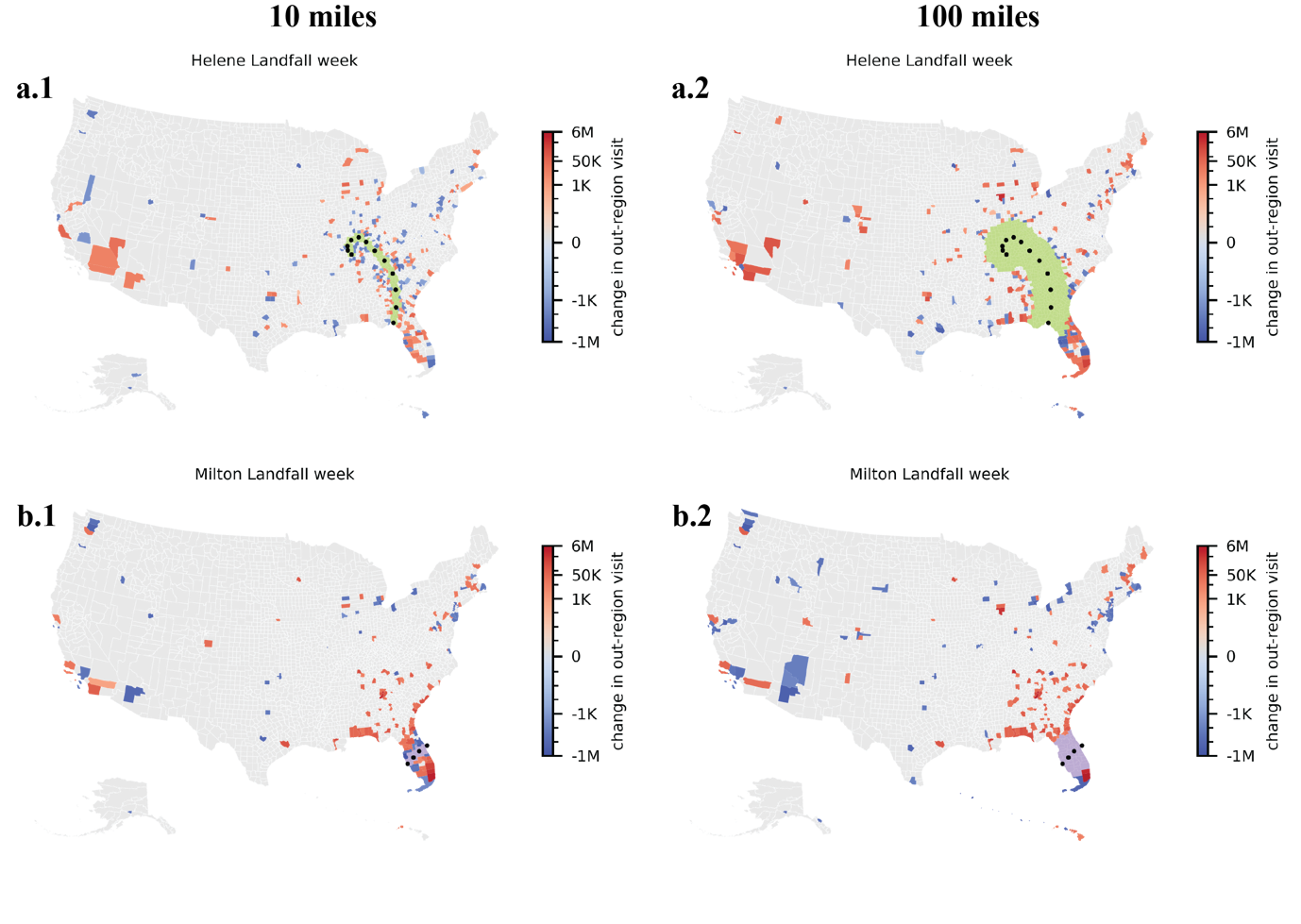

**Fig.S8 Changes in out-region visits across destination counties at different distance thresholds.** Colored counties indicate statistically significant increases or decreases in out-region visits relative to natural mobility variation. Panels **a.1** and **a.2** show results for Hurricane Helene at 10-mile and 100-mile thresholds, respectively. Panels **b.1** and **b.2** depict results for Hurricane Milton at the same thresholds.

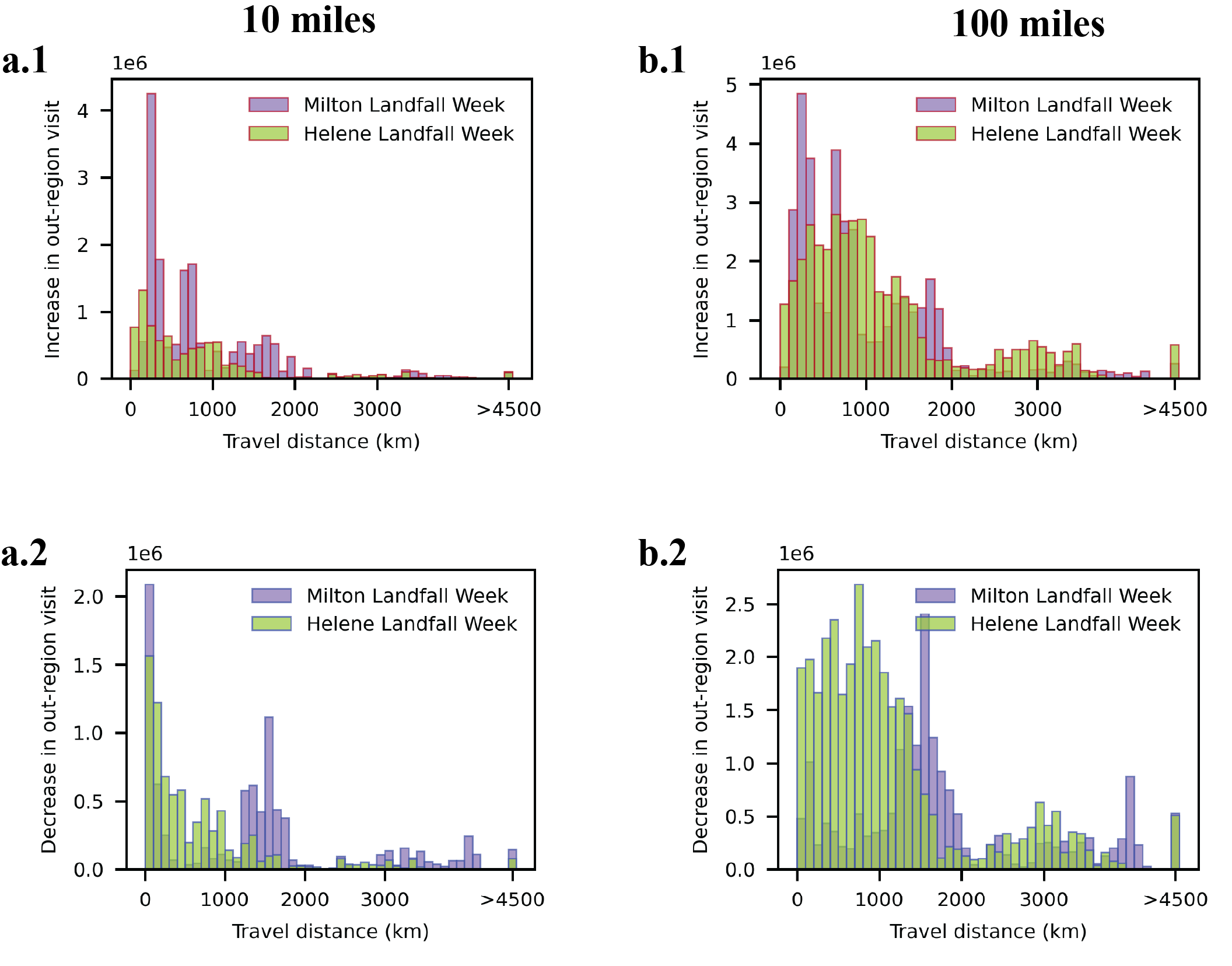

**Fig.S9 Distribution of visit changes across various travel distances.** The histograms show the distribution of out-region visit changes to destination counties, grouped by travel distance. Panels **a** and **b** correspond to affected regions defined by 10-mile and 100-mile thresholds, respectively. For Hurricane Helene, the average weighted travel distances are 463.2 km (10-mile) and 883.4 km (100-mile). For Hurricane Milton, the respective averages are 774.0 km and 1002.3 km.

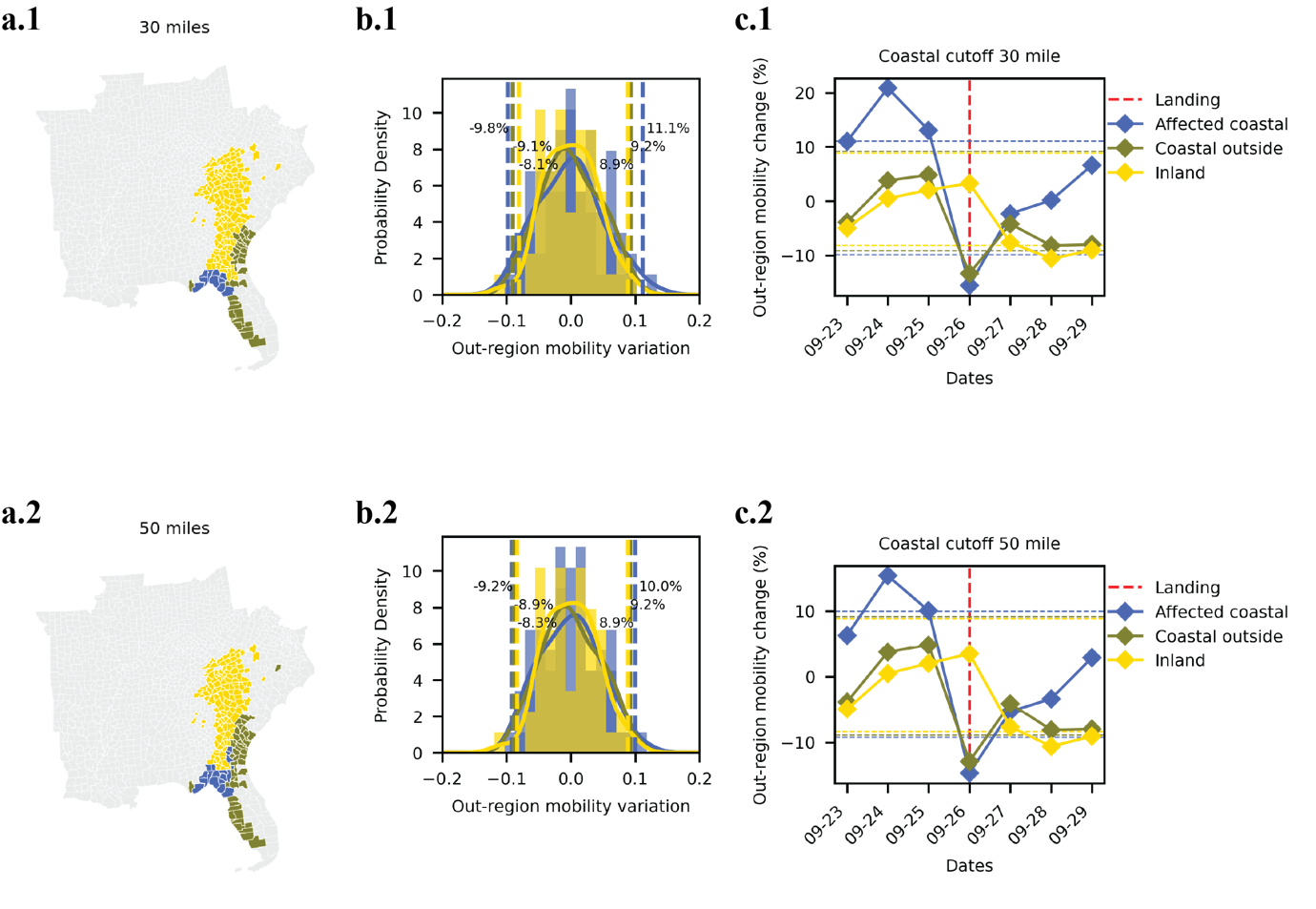

**Fig. S10. Out-region Mobility Variation in FEMA-Declared Disaster Counties During Hurricane Helene.** Panels a.1 and a.2 classify counties designated as FEMA Major Disaster Areas using two coastal proximity thresholds: 30 miles (a.1) and 50 miles (a.2). Counties are grouped into: (i) coastal counties within both the coastal proximity threshold and 50 miles of the storm track (blue), (ii) coastal counties outside the 50-mile storm track cutoff (olive), and (iii) inland counties (yellow). Panels b show the distributions of out-region mobility variation for each group in the same period last year (July 15^th^, 2023, to Sep 9^th^, 2023), with vertical dashed lines indicating the 95% confidence intervals of natural variation. Panels c present the time series of daily out-region mobility changes (%), with red dashed lines marking the hurricane landfall date.

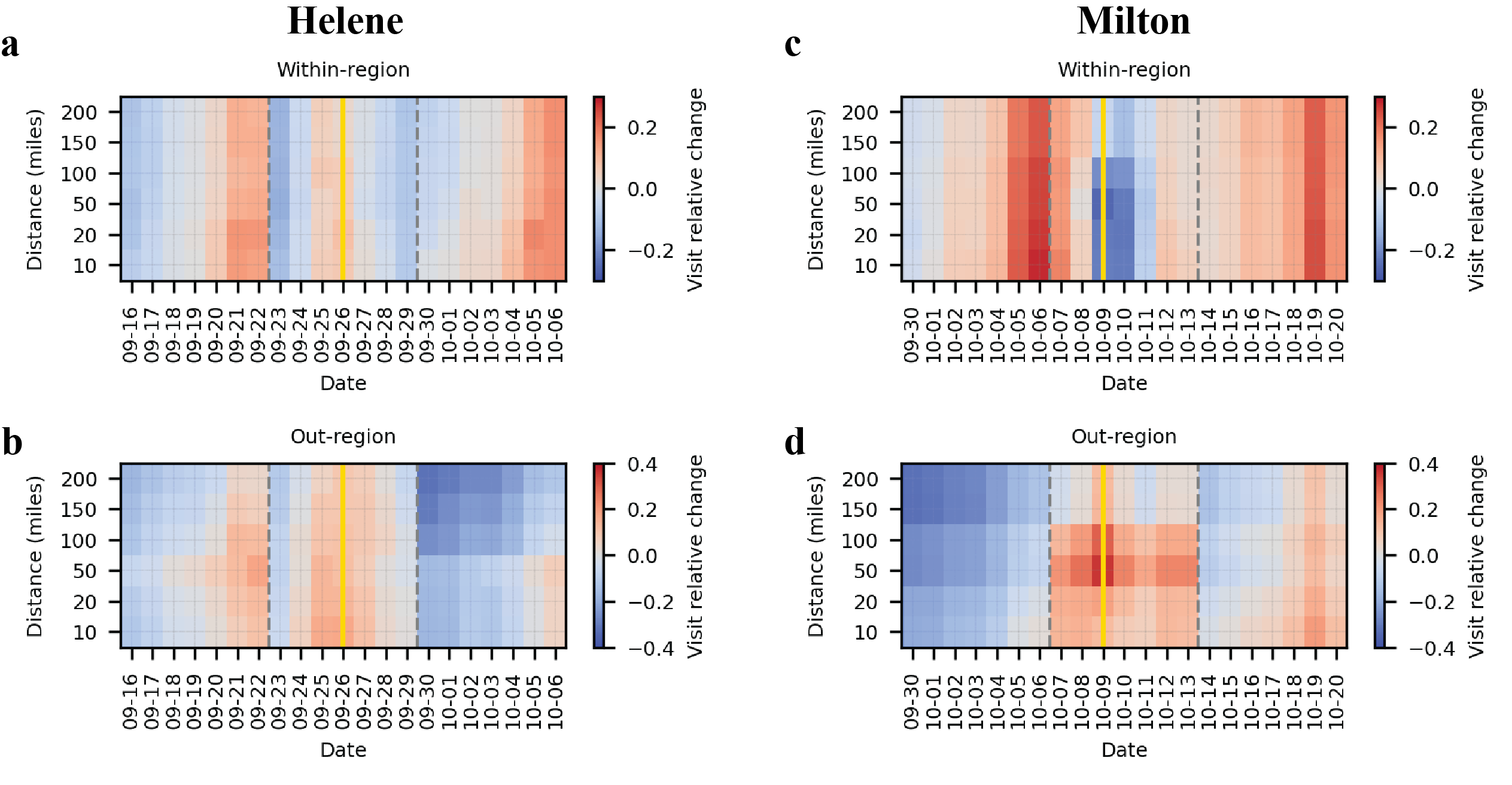

Fig. S11. Relative changes in within-region (top panels) and out-region (bottom panels) mobility when using the week of August 5th–11th, 2024 as the baseline, instead of September 9th–15th, 2024 as in the main analysis. The yellow vertical lines indicate the hurricane landfall dates, and dashed grey lines mark the landfall weeks. Each row represents the affected region defined using different distance thresholds to the storm track. Results show that the overall patterns remain consistent with our main findings.

**Tab.S1 Number of affected counties, destination counties and population for different distances.**

| **Cut-off (miles)** | **No. affected counties** | | **No. destination counties** | | **Population** | |
| --- | --- | --- | --- | --- | --- | --- |
|  | Helene | Milton | Helene | Milton | Helene | Milton |
| 10 | 99 | 8 | 437 | 218 | 3,744,311 | 5,399,152 |
| 50 | 271 | 21 | 230 | 310 | 14,533,220 | 9,889,433 |
| 100 | 487 | 34 | 341 | 286 | 28,866,947 | 13,571,589 |

|  | State | Wind | Storm Surge | Freshwater Flooding | Tornado | Falling Trees | Total |
| --- | --- | --- | --- | --- | --- | --- | --- |
| Helene | Florida | 4 | 14 |  |  |  | 18 |
|  | Georgia | 26 |  |  | 2 |  | 28 |
|  | South Carolina | 24 |  | 2 |  |  | 26 |
|  | North Carolina | 8 |  | 78 |  |  | 86 |
|  | Tennessee |  |  | 15 |  |  | 15 |
|  | Virginia | 2 |  |  |  |  | 2 |
|  | Indiana | 1 |  |  |  |  | 1 |
|  | **Total** | 65 | 14 | 95 | 2 |  | 176 |
| Milton | Florida |  |  | 2 | 6 | 4 | 12 |
|  | **Total** |  |  | 2 | 6 | 4 | 12 |

Tab. S2. Direct deaths by state and cause for Hurricanes Helene and Milton, adapted from NOAA’s Tropical Cyclone Reports [1,2]. These data highlight geographic differences in dominant mortality risks, with freshwater flooding dominating in North Carolina, wind in Georgia and South Carolina, and storm surge and tornado in Florida. There are 15 deaths known due to Milton—12 in the United States as shown in the table and 3 in Mexico.

[1] National Hurricane Center. Tropical Cyclone Report: Hurricane Helene (AL092024). NOAA (2024). Available at: https://www.nhc.noaa.gov/data/tcr/AL092024_Helene.pdf

[2] National Hurricane Center. Tropical Cyclone Report: Hurricane Milton (AL152024). NOAA (2024). Available at: https://www.nhc.noaa.gov/data/tcr/AL152024_Milton.pdf
